# SARS-CoV-2 susceptibility and *ACE2* gene variations within diverse ethnic backgrounds

**DOI:** 10.1101/2021.08.18.21261804

**Authors:** Nirmal Vadgama, Alexander Kreymerman, Jackie Campbell, Olga Shamardina, Christiane Brugger, Genomics England Research Consortium, Richard T. Lee, Christopher J. Penkett, Casey A. Gifford, Mark Mercola, Jamal Nasir, Ioannis Karakikes

## Abstract

**Background:** Host genetics play a major role in COVID-19 susceptibility and severity. Here, we analyse an ethnically diverse cohort of National Health Service (NHS) patients in the United Kingdom (UK) to assess the association between variants in the *ACE2* locus and COVID-19 risk.

**Methods:** We analysed whole-genome sequencing (WGS) data of 6,274 participants who were tested for SARS-CoV-2 from the UK’s 100,000 Genomes Project (100KGP) for the presence of *ACE2* coding variants and expression quantitative trait loci (eQTLs).

**Findings:** We identified a splice site variant (rs2285666) associated with increased ACE2 expression with an overrepresentation in SARS-CoV-2 positive patients relative to 100KGP controls (p = .015), and in hospitalised European patients relative to outpatients in intra-ethnic comparisons (p = .029). We also compared the prevalence of 288 eQTLs, of which 23 were enriched in SARS-CoV-2 positive patients. The eQTL rs12006793 had the largest effect size (d = 0.91), which decreases ACE2 expression and is more prevalent in controls, thus potentially reducing risk of COVID-19. We identified three novel nonsynonymous variants predicted to alter ACE2 function, and showed that three variants (p.K26R, p.H378R, p.Y515N) alter receptor affinity for the viral Spike (S) protein. Variants p.K26R and p.N720D are more prevalent in the European population (p < .001), but Y497H is less prevalent compared to East Asians (p = .020).

**Interpretation:** Our results demonstrate that the spectrum of genetic variants in *ACE2* may inform risk stratification of COVID-19 patients and could partially explain the differences in disease susceptibility and severity among different ethnic groups.

**Funding:** The 100KGP is funded by the National Institute for Health Research and NHS England. Funding was also obtained from Stanford University, Palo Alto.

## INTRODUCTION

Severe acute respiratory syndrome-coronavirus 2 (SARS-CoV-2) is a novel positive-strand RNA virus identified as the cause of the coronavirus disease 2019 (COVID-19). It is responsible for a pandemic that has cost the lives of over 3.84M people since the first documented case in Wŭ hàn, China, in December 2019.^1^

Disproportionate hospitalisations, deaths, and complications from SARS-CoV-2 in some minority groups suggests there might be a potential biological underpinning driving risk disparities in COVID-19 patients. Although many health ministries have identified age and chronic conditions, such as obesity, diabetes, cancer, and immunodeficiency, as major drivers of risk, genetic markers of susceptibility have not yet been used as a metric for risk.^2^

The most well-known and evaluated mechanism of SARS-CoV-2 infection is the binding and uptake of viral particle through the human angiotensin-converting enzyme 2 (ACE2) receptor, a type 1 integral membrane glycoprotein^3^. This process is initiated by the trimeric transmembrane spike (S) glycoprotein protruding from the viral surface. During infection, the S protein is cleaved into subunits, S1 and S2. S1 contains the receptor binding domain (RBD), which allows the virus to directly bind to the peptidase domain of ACE2, while S2 is responsible for membrane fusion upon viral infection.^1,4^ Thus, coding variants within *ACE2* could have the potential to alter SARS-CoV-2 binding and possible infection responses in different individuals based on host genetics. Host genetics is known to play an important role in susceptibility to other viral infectious diseases, including SARS-CoV, HIV and influenza.^5^ In addition, ACE2 expression has been shown to vary between different populations, driven in part by expression quantitative trait loci (eQTLs).^6^

Several studies have shown that both *ACE2* coding variants and eQTLs can influence the host susceptibility or resistance to SARS-CoV-2 infection, by altering S protein binding or ACE2 expression. However, many of these studies are based on *in silico* predictions from public datasets or have relatively few SARS-CoV-2 positive patient sample sizes, of which there is often limited diversity.^7–9^

To identify population level variants that might contribute to SARS-CoV-2 infection differences, we analysed *ACE2* variants within the 100,000 Genomes Project (100KGP). This dataset consists of whole-genome sequencing (WGS) data from patients recruited through the National Health Service (NHS) representing the diverse population of the United Kingdom (UK)^10^, including 1,837 SARS-CoV-2 positive patients and 37,207 unrelated individuals with no record of SARS-CoV-2 infection, aggregated by five continental ancestries. Using this dataset, we provide evidence that *ACE2* variants play a role in COVID-19 susceptibility and severity, with population specific effects.

Our findings are geared towards establishing a structure-function predictive framework for exploring genotype-phenotype correlations, and a genetic trait score to estimate individual liability to disease.

## MATERIALS AND METHODS

### Patient subjects

The 100KGP was established to sequence 100,000 genomes from around 85,000 NHS patients in the UK affected by a rare disease, cancer, or infectious disease, with detailed phenotype and clinical data (https://www.genomicsengland.co.uk/).^10^ Combining WGS data with medical records has created a ground-breaking research resource enriched for genetic diseases. As part of its recruitment target, Genomics England has highlighted the importance of engaging with the UK’s increasingly diverse black and minority ethnic population, who are underrepresented in clinical studies and research.

This dataset consisted of 6,274 patients who were tested for SARS-CoV-2, of which 1,837 tested positive. Of the positive cases, 1,411 (77%) were classified as not hospitalised, and 426 (23%) were classified as inpatients or deceased. We also used WGS data from a large control group consisting of 37,207 unrelated individuals from 100KGP (males = 17,066; females = 20,141; total allele number = 57,348), to scan for *ACE2* variants and eQTL distribution in different populations.

The study adheres to the principles set out in the Declaration of Helsinki. Patients and relatives gave written informed consent for genetic testing. Ethical approval for The 100,000 Genomes Project was granted by the East of England - Cambridge South Research Ethics Committee (REC Ref 14/EE/1112).

### Ancestry inference

Using multi-sample VCFs for participants in the 100KGP cohort, we generated Principal Components (PCs), calculated pairwise relatedness among samples, and estimated probabilities of genetic ancestry for five broad superpopulations. Broad genetic ancestry was estimated using ethnicities from the 1000 genomes project phase 3 (1KGP3), by generating PCs for 1KGP3 samples and projecting all 100KGP participants onto these. The five broad superpopulations are European, South Asian, African, East Asian, Ad Mixed American, as well as multiracial individuals, labelled as Other.

### Classification of hospitalised status

Patient hospitalised status is based on whether the collected specimen was from an acute (emergency) care provider, an accident and emergency department, an inpatient location, or resulted from health care associate infection. ‘Not hospitalised’ is interpreted as there being no evidence in the microbiological record that the patient has been an inpatient, but this may still be a possibility as the microbiology data source is not linked to admissions data.

### Computational modelling of ACE2

We employed a computational method to predict the effect of the coding variants identified in the 100KGP dataset, based on homology modelling and computational docking of the virus-receptor protein-protein interaction. Several crystal structures of the SARS-CoV-2 RBD and ACE2 have been reported (pdb identifiers: 6VW1,^11^ 6LZG,^12^ 6M0J,^4^ 7A94,^13^) allowing us to demonstrate the utility of this model. Amino acids of interest were mutated in Coot^14^ and visually analysed using UCSF Chimera.^15^ Mutations were ranked according to their potential to affect protein stability and activity. Structure figures were prepared in Chimera.

### Public gene expression dataset acquisition and analysis

We obtained expression data through the genotype-tissue expression (GTEx) database (https://gtexportal.org/home/).^16^ This data was also used to extract quantitative trait loci (eQTLs) for ACE2 (all data based on RNA-seq experiments). Population allele frequencies were obtained from 100KGP.

### Coding variant annotation and function prediction

The raw list of SNVs and indels among cases and controls were annotated using ANNOVAR.^17^ Variants in splicing regions, 5′-UTR, 3′-UTR, and protein-coding regions, such as missense, frameshift, stop loss, and stop gain mutations, were considered. *In silico* prediction of pathogenicity was assessed using PolyPhen2,^18^ CADD,^19^ REVEL^20^ and GERP++^21^ scores. Highest priority variants were nonsynonymous or in the splice site, with a corresponding REVEL score above the default threshold of 0.5, a GERP++ score greater than two, and PolyPhen2 prediction outcome of ‘probably damaging’.

### Statistical analysis

The data was analysed using chi-square tests (with Fisher’s exact where any expected value < 5), and Z-tests for group differences (with Bonferroni correction). Groups with no observed counts were excluded from the analysis. Statistical significance was taken at 5% level (two-tailed).

## RESULTS

### Summary of SARS-CoV-2 positive cases

To see if there is an observable difference in SARS-CoV-2 positivity and disease severity in different ethnic backgrounds, we used a unique dataset containing genetic information within the 100KGP. This dataset contains the SARS-CoV-2 test results, genetically predicted ancestry, and hospitalisation status of each patient. We stratified positive cases in the 100KGP by the five predominant superpopulations representing the general population in UK **(Figure 1A)**. Our data shows that 1,254 individuals of European (68.3%), 322 of South Asian (17.5%), 49 of African (2.7%), 15 of East Asian (0.8%), 4 of Ad Mixed American (0.2%) background, as well as 193 uncategorised individuals, labelled as Other (10.5%), tested positive for SARS-CoV-2 **(Figure 1B)**.

**Figure 1:**
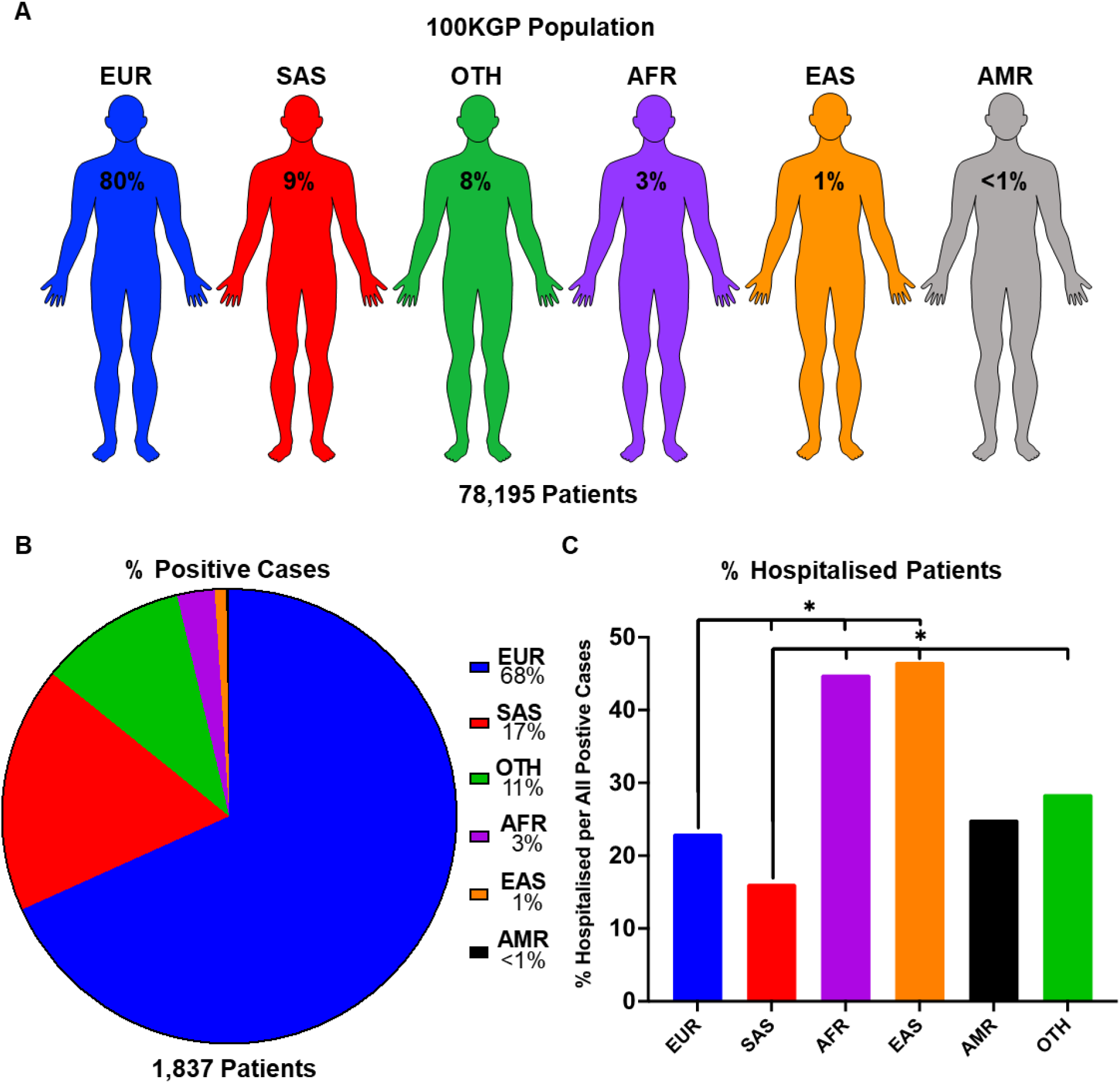
SARS-CoV-2 infection numbers and hospitalisation rate per ethnic group in the UK population. (**a**) The proportion of individuals in each population of the entire 100KGP cohort assigned with a probability of > 0.8 for any one ancestry. (**b**) Percentage of patients who tested positive for SARS-CoV-2, presented as positive cases in each ethnic background relative to the total positive cases in the 100KGP. (**c**) Percentage of hospitalised patients per total positive individuals in each ethnic group. P values are calculated using Chi-square test.

The chi-square test was used to compare proportions of inpatients, demonstrating that hospitalised outcomes in SARS-CoV-2 positive patients were statistically different across ethnic groups (χ^2^ = 129.64, df = 5, *p* < 0.001). Individuals of East Asian and African background were approximately 50% more likely to be hospitalised, as compared to Europeans. Furthermore, the largest difference in hospitalisation rate was found between South Asians (16.2% hospitalised) and East Asians (46.8% hospitalised) **(Figure 1C; Table S1)**, suggesting that individuals of South Asian decent are less likely to be hospitalised from SARS-CoV-2 infections. In addition, Ad Mixed Americans did not statistically differ between any ethnic group, which may be due to a small sample size (*n* = 4). Nonetheless, these data provide evidence for varying degrees of susceptibility to COVID-19 in different ethnicities.

We hypothesise that these findings may reflect an underlying biological driver for viral susceptibility in different genetic backgrounds. To investigate this, we interrogated the 100KGP patient database for correlations between SARS-CoV-2 positivity status and genetic variants that alter *ACE2* protein structure, as well as expression. Although there are many mechanisms involved in SARS-CoV-2 entry and host responses, we focused on ACE2 because it is the main host receptor for viral entry and a limiting factor for infection.

### ACE2 coding variants in 100KGP controls

To identify whether variants within *ACE2* could have a predicted influence over S protein-host cell interactions, we first identified coding variants within 37,207 unrelated individuals in the 100KGP dataset. This dataset represents the ethnic diversity of the UK population. Coding variants were stratified by the five predominant superpopulations represented within the UK population (European, South Asian, African, Ad Mixed American, and East Asian descent) and by allelic frequency. The resulting analysis provided 114 SNVs **(Figure 2A)**, three deletions, and one insertion leading to a frameshift identified in one individual of European descent **(Table S2)**.

**Figure 2:**
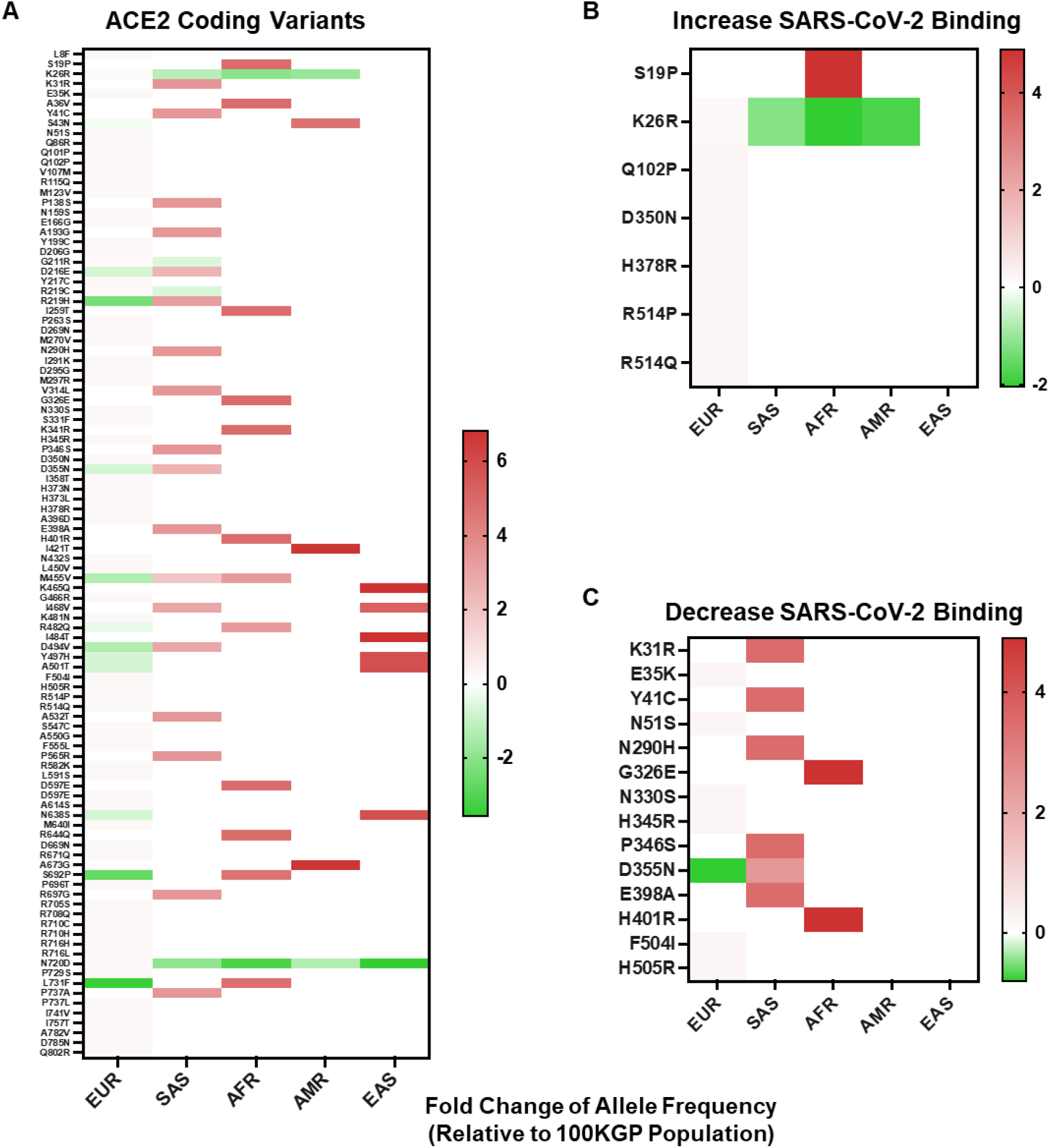
Allelic frequency of nonsynonymous *ACE2* variants in different ethnicities. (**a**) Allelic frequency of *ACE2* coding variants present in individual ethnicities, graphed as a log base 2-fold change relative to the allelic frequency in 100KGP unrelated controls (n = 37,207). (**b)** Allelic frequency of *ACE2* coding variants in individual ethnicities that have been shown to increase SARS-CoV-2 S protein binding affinity for ACE2. (**c**) Allelic frequency of *ACE2* coding variants in individual ethnicities that have been shown to decrease SARS-CoV-2 S protein binding affinity for ACE2. All heat maps are graphed with values below and above zero set relative to the dataset. Shades of red indicate increased allelic frequency and green decreased allelic frequency, with lighter shades appearing as values move closer to no appreciated change relative to the 100KGP allelic frequencies, as indicated by white. Coding variants displayed by protein location and amino acid change.

To see if any of these variants have the potential to effect binding of S protein, we compared our findings against data from a high-throughput functional mutagenesis screen that used deep sequencing to assess S-protein binding abilities to 2340 ACE2 variants.^22^ In the 100KGP control dataset, we identified 21 *ACE2* nonsynonymous variants that alter S protein binding, seven that increase binding and 14 decrease binding **(Figure 2B and C)**. We stratified these variants based on allelic frequency within different ethnic backgrounds. Populations of African and South Asian descent generally had the greatest number of alleles with increased frequency, relative to the general population **(Figure 2A)**. This was also reflected in variants that both increase and decrease S protein binding affinity for ACE2 **(Figure 2B and C)**. Specifically, we detected a greater frequency of variants in South Asians that decrease SARS-CoV-2 S protein binding affinity for ACE2 **(Figure 2C)**, and one variant enriched within African descent populations that increases S protein-binding affinity for ACE2 **(Figure 2B)**.

### ACE2 coding variants in SARS-CoV-2 positive patients

To see if any coding variants in ACE2 correlate with decreased or increased propensity for SARS-CoV-2 positivity, we analysed WGS data of 1,837 SARS-CoV-2 positive patients and identified 18 nonsynonymous *ACE2* variants **(Table 1; Table S3)**. Six nonsynonymous variants were novel (p.V316L, p.H345R, p.H401R, p.M455V, p.Y515N, p.P737L), of which three are predicted to be deleterious based on *in silico* predictions. Three variants (p.K26R, p.H378R, p.Y515N) found across the interaction surface were shown to alter receptor affinity for the viral S protein based on deep mutagenesis experiments.^22^ Variants p.H378R and p.K26R were found to increase binding, whereas the novel p.Y515N variant decreased binding.

**Table 1:** Characterisation of nonsynonymous ACE2 variants in SARS-CoV-2 patients. Intronic, intergenic and synonymous variants are considered non-pathogenic and therefore excluded from the list. The index score is based on the type of variant, the REVEL score, and the GERP++ score. An index score of 1 signifies a nonsynonymous variant with corresponding REVEL score < 0.5, suggesting low pathogenicity. An index score of 2 indicates nonsynonymous variant with a REVEL score of > 0.5, GERP++ score > 2, CADD score of > 20 and PolyPhen2 prediction outcome of ‘probably damaging’, suggesting high pathogenicity. Six variants were not reported in public databases (p.V316L, p.H345R, p.H401R, p.M455V, p.P737L, p.Y515N). Deep mutagenesis experimental data show p.K26R, p.H378R and p.Y515N altered binding affinity with S protein.^22^ This is consistent with our protein interaction analysis. Mutations are ranked according to their potential to affect protein stability and activity, denoted by asterisks (* = low, ** = medium, *** = high).

| Variant | rsID | AA Change | Index | Protein modelling | S protein binding affinity in RBD | Minor allele frequency within 100KGP |  |  |  |  |  |
| --- | --- | --- | --- | --- | --- | --- | --- | --- | --- | --- | --- |
|  |  |  |  |  |  | Combined | EUR | SAS | AFR | AMR | EAS |
| chrX:15564123G>A | novel | p.P737L | 1 | - | NA | 1.74E-05 | 2.03E-05 | 0 | 0 | 0 | 0 |
| chrX:15564142G>A | rs147311723 | p.L731F | 1 | - | NA | 6.63E-04 | 6.09E-05 | 0 | 1.81E-02 | 0 | 0 |
| chrX:15564175T>C | rs41303171 | p.N720D | 1 | - | NA | 2.34E-02 | 2.64E-02 | 6.09E-03 | 2.58E-03 | 9.21E-03 | 1.98E-03 |
| chrX:15566293A>G | rs149039346 | p.S692P | 2 | - | NA | 4.06E-05 | 0 | 0 | 6.71E-03 | 0 | 0 |
| chrX:15572271C>T | rs763593286 | p.A532T | 1 | * | NA | 5.23E-05 | 0 | 5.89E-04 | 0 | 0 | 0 |
| chrX:15572322A>T | novel | p.Y515N | 2 | ** | Decrease | 0 | 0 | 0 | 0 | 0 | 0 |
| chrX:15573419A>G | rs961921482 | p.Y497H | 1 | * | NA | 3.49E-05 | 2.03E-05 | 0 | 0 | 0 | 1.98E-03 |
| chrX:15575745T>C | novel | p.M455V | 1 | ** | NA | 5.23E-05 | 2.03E-05 | 1.96E-04 | 5.17E-04 | 0 | 0 |
| chrX:15578124A>G | rs1352508510 | p.I421T | 1 | * | NA | 1.74E-05 | 0 | 0 | 0 | 1.84E-03 | 0 |
| chrX:15578184T>C | novel | p.H401R | 2 | ** | Negligible | 1.74E-05 | 0 | 0 | 5.17E-4 | 0 | 0 |
| chrX:15578253T>C | rs142984500 | p.H378R | 2 | *** | Increase | 1.92E-04 | 2.23E-04 | 0 | 0 | 0 | 0 |
| chrX:15581257T>C | novel | p.H345R | 2 | ** | Negligible | 1.74E-05 | 2.03E-05 | 0 | 0 | 0 | 0 |
| chrX:15581269T>C | rs138390800 | p.K341R | 1 | * | NA | 2.27E-04 | 0 | 0 | 6.71E-03 | 0 | 0 |
| chrX:15581345C>A | novel | p.V316L | 1 | * | NA | 1.74E-05 | 0 | 1.96E-04 | 0 | 0 | 0 |
| chrX:15587768G>A | rs200745906 | p.P263S | 2 | * | NA | 1.92E-04 | 2.23E-04 | 0 | 0 | 0 | 0 |
| chrX:15589385G>A | rs372272603 | p.R219C | 1 | * | NA | 9.42E-04 | 1.04E-03 | 5.89E-04 | 0 | 0 | 0 |
| chrX:15589409C>T | rs148771870 | p.G211R | 1 | * | NA | 2.14E-03 | 2.35E-03 | 1.37E-03 | 0 | 0 | 0 |
| chrX:15600835T>C | rs4646116 | p.K26R | 1 | * | Increase | 6.42E-03 | 7.10E-03 | 2.75E-03 | 1.55E-03 | 1.84E-03 | 0 |

In addition, we analysed the effect of these variants on protein structure and assessed if they might alter the interaction between ACE2 and the SARS-CoV-2 S protein **(Figure 3)**. While predicting the effect of single mutations in the core of the protein on the interaction of ACE2 with the S protein are challenging, we can nevertheless make predictions on the effect on protein stability. We picked five mutations for structural analysis (H345R, H378R, H401R, M455V and Y515N). H378R is the mutation with the most severe destabilisation effect on ACE2 as it disrupts a Zn binding motif. Similarly, H401R disrupts an H-bond between H378 and H401, destabilising the motif, albeit to a lesser extent. H345R does not interact directly with either of the amino acids that coordinate the Zn ion, but it is in close proximity and could have a negative effect. Y515N can be found lining the pocket providing access to the Zn motif and mutations could interfere with potential interaction partners. S-aromatic motifs have been reported to have stabilising effects on proteins,^23^ thus substituting methionine for valine at position 455 can have a detrimental effect on protein stability.

**Figure 3:**
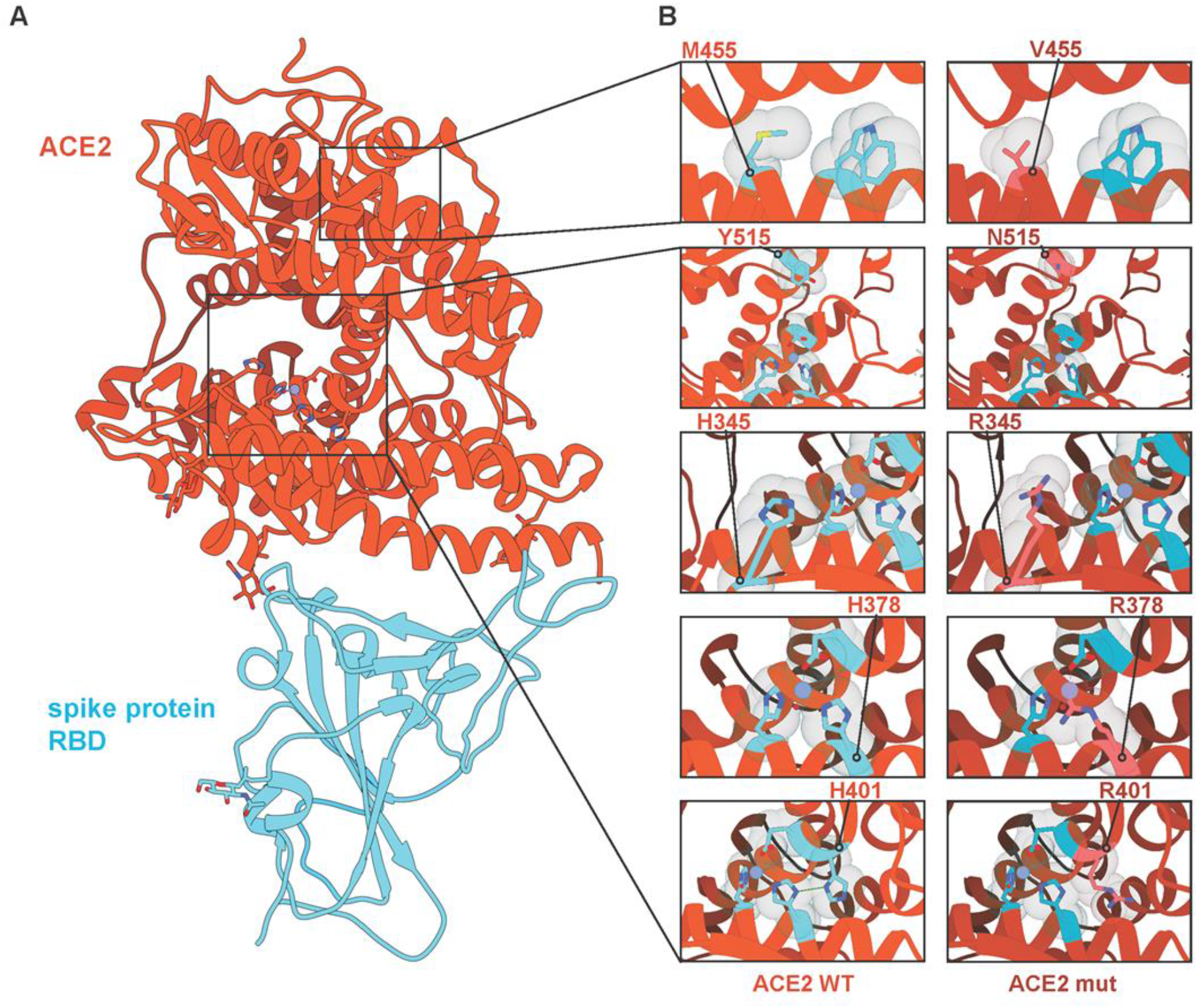
Interaction diagrams of *ACE2* variants and SARS-CoV-2 S-protein. (**a**) Crystal structure of ACE2 and SARS-Cov-2 S protein (pdb identifier: 6LZG) with ACE2 coloured in red-orange and the RBD domain of the SARS-CoV-2 S protein in light blue. Regions of interest are highlighted and expanded in panel (**b**), showing mutated residues M455V, Y515N, H345R, H378R and H401R with wild-type sequences (left) and mutated sequences (right). Hydrogen bonds are shown in green, Zinc ions are shown in purple and water molecules are shown in red.

### ACE2 splice variants in SARS-CoV-2 positive patients

We also identified an *ACE2* splice region variant (rs2285666 C/T), which was significantly more prevalent among SARS-CoV-2 patients compared to the control population (*p* = .015). Further, intra-ethnic comparisons showed that this variant was more prevalent in inpatients (minor allele frequency (MAF) = 23.9%) compared to patients not hospitalised (MAF = 18.4%) within the European population (*p* = .029), suggesting it plays a role in susceptibility and severity **(Table S4)**.

The rs2285666 C/T SNP is located on the fourth base position of intron three, which may affect gene expression via mRNA alternate splicing mechanisms.^24^ According to HaploReg tool (v4.1), rs2285666 alters transcription factor binding sites; namely, HNF1, and Ncx motifs. Using an ELISA method, Wu et al.^25^ showed that the rs2285666 SNP elevated serum ACE2 levels, with the T/T genotype increasing expression by almost 50%. Similarly, the GTEx database classifies this variant as a significant eQTL, which is associated with an increased expression of ACE2, CA5BP1, PIR and TMEM27, and decreased expression of CA5B.

Of note, previous studies have identified rs2285666 as a risk factor for hypertension, type 2 diabetes, and coronary heart disease.^26,27^ Taken together, this polymorphism may increase SARS-CoV-2 infection rates by altering ACE2 expression as well as predisposing to comorbidities observed in COVID-19 patients.

We also analysed *ACE2* variants in the 3′UTR, 5′UTR and promoter regions, where the promoter is defined as 2 kb upstream of the start codon. No significant differences in the allele frequencies were found between SARS-CoV-2 positive patients and controls, or patients stratified by hospitalised status. However, when examining intra-ethnic differences in allele and haplotype frequencies, the rs199651576 T/C 5’UTR polymorphism was found to be higher in SARS-CoV-2 patients (MAF = 0.12%) compared to controls (MAF = 0.016%) within the European population (P=0.039) **(Table S4)**. The function of this variant remains unclear, although it is possible that it is involved with the translational regulation of the ACE2 mRNA transcript.

### Inter-ethnic differences in ACE2 coding variants

To see if any of the variants found within SARS-CoV-2 positive patients differs between ethnicities, we compared the MAFs from 37,207 unrelated individuals, representing the general population of the UK. Z-test comparisons were made between ethnic groups (with Bonferroni correction where there were more than two groups), and the chi-square test was used to see whether distributions of variables differ from each another. Variants p.K26R (χ^2^ = 23.12, df = 3, *p* < 0.001) and p.N720D (χ^2^ = 137.55, df = 4, *p* < 0.001) were statistically different between groups. These variants were also the most frequent in the 100KGP cohort and Genome Aggregation Database (gnomAD).^28^

The p.K26R variant was more common in Europeans (MAF = 0.710%) compared to South Asians (MAF = 0.275%; *p* < .001) and Africans (MAF = 0.155%; *p* = .001). The K26 residue lies on the proximal end of the SARS-CoV-2 RBD–ACE2 interface.^4^ Structural predictions have shown that the K26R variant enhances the affinity of ACE2 for SARS-CoV-2 by strengthening the hydrogen bond between H34 and Y453 of the S-protein.^29,30^ This is consistent with deep mutagenesis experimental data using a synthetic human ACE2 mutant library.^22^

Variant p.N720D was also more common in Europeans (MAF = 2.64%) compared to South Asians (MAF = 0.609; *p* < .001), Africans (MAF = 0.285%; *p* < .001) and East Asians (MAF = 0.198; *p* < .001). The p.N720D variant lies in the C-terminal collectrin domain of ACE2. Although it is not involved in the SARS-CoV-2 S-protein interaction, studies have shown that it enhances transmembrane protease, serine 2 (TMPRSS2) binding and cleavage of ACE2. As TMPRSS2 cleavage of the ACE2 receptor increases viral entry, the p.N720D variant may lead to increased COVID-19 susceptibility.^29,31^

Finally, the p.Y497H variant was more prevalent in East Asians (MAF = 0.198%) compared to Europeans (MAF = 0.00203%) (*p* < .001), but absent in other populations **(Table 1; Table S5)**.

### ACE2 eQTLs in SARS-CoV-2 positive patients

While protein-protein interactions can explain some components of viral entry and potential susceptibility in variant carrying at-risk groups, differential expression of ACE2 may also be a contributor to viral-host cell interactions.

Among the 31 GTEx human tissues, the highest ACE2 expression levels are found in the testis, small intestine, kidneys, heart, thyroid, and adipose tissue.^16^ The lowest ACE2 expression levels are found in whole blood, spleen, brain, and skeletal muscle. The lungs, pancreas, oesophagus (muscularis) and liver had intermediate ACE2 expression levels.

The Human Protein Atlas (HPA)^32^ database shows that ACE2 had relatively high expression levels in the duodenum, small intestine, gallbladder, kidneys, testis, seminal vesicle, colon, rectum, and adrenal gland. The HPA database also showed that the gastrointestinal tract (duodenum, small intestine, colon, and rectum), kidney, gallbladder, and male tissues (testis and seminal vesicle) had high expression levels of both the ACE2 protein and gene. Interestingly, the results obtained from the HPA (fresh frozen tissue) and GTEx (postmortem tissue) are similar, suggesting negligible effects of the sampling procedures used by the GTEx consortium on RNA degradation.

We also interrogated the GTEx database for the distribution of ACE2 eQTLs (GTEx analysis Release V8). For ACE2, we identified 288 significant (FDR < 0.05) eQTLs in 20 different tissues, including, adipose (*n* = 31; 10.8%), brain (*n* = 246; 85.4%), testis (*n* = 2; 0.7%), and prostate (*n* = 60; 20.8%). It would be highly valuable to compare the frequency of eQTL variants with ACE2 expression specifically in the lung with susceptibility to viral infection and severity of COVID-19. However, to date, no eQTL for ACE2 in the lung has been reported in the GTEx database, thus warranting further investigations in this regard.

The allele frequencies of the 288 eQTL variants were compared among different ethnicities in the 100KGP dataset **(Figure 4; Table S6)**. Across this cohort, we performed a case-control genetic association analysis on ACE2 eQTLs. Of the 288 eQTLs associated with tissue expression of ACE2 in the GTEx database, 132 were significant between patients and controls. Twenty-three were significantly different after Bonferroni correction, seven of which had a Cohen’s effect size, d, of > 0.2. The largest MAF difference was observed in rs12006793 (d = 0.91). This eQTL decreases ACE2 expression and was more common in the control group **(Tables S7 and S9)**, suggesting it reduces COVID-19 risk. This eQTL was statistically different between ethnic groups (χ^2^ = 801.06, df = 4, *p* < 0.001). The eQTL prevalence significantly differed between all ethnic comparisons, except between Ad Mixed Americans and East Asians, and Ad Mixed Americans and Europeans. It was more common in Africans (MAF = 66.6%) and South Asians (MAF = 60.1%). However, according to GTEx, it was only expressed in adipose tissue (omentum) with a normalised enrichment score of -0.1.

**Figure 4:**
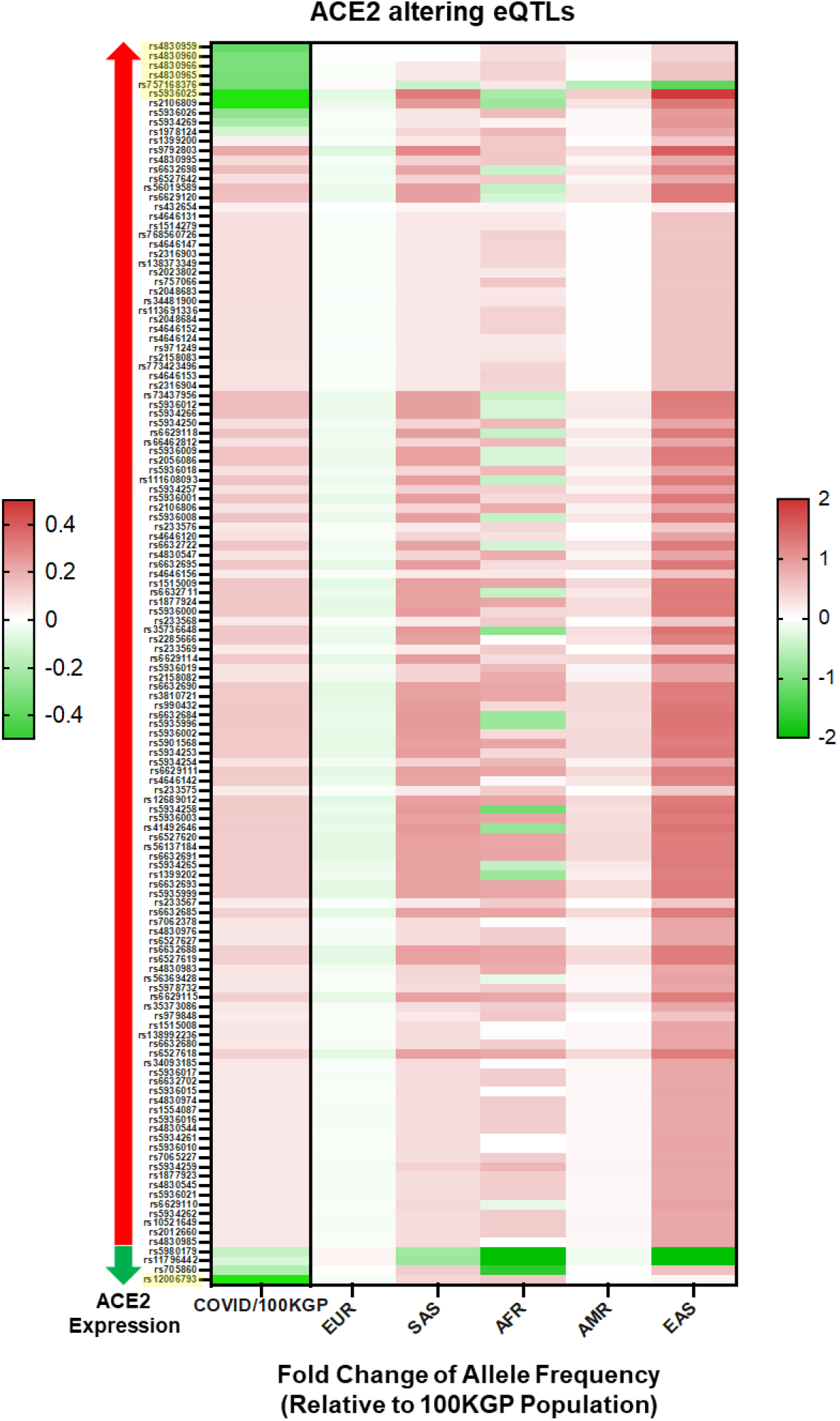
Statistically significant eQTLs in SARS-CoV-2 positive individuals. Displayed by total positive individuals (value on the left) and by ethnic background (value on the right), values are presented as log base 2-fold change relative to the allelic frequency within non-positive individuals found in the 100KGP (n = 37,207). eQTLs are also subdivided by effects on a ACE2 expression, with eQTLs known to associate with increased ACE2 expression marked by a red arrow and those associated with decreased ACE2 expression marked with a green arrow. All displayed eQTLs are significant based on chi-squared test, and yellow highlighted eQTLs had Cohen’s values above 0.2. All eQTLs are referenced by rs identifiers.

When comparing hospitalised SARS-CoV-2 positive patients with patients not hospitalised, only two eQTLs were statistically significant after multiple testing correction, but none had a Cohen’s effect size greater than 0.2. Interestingly, however, rs12006793 was one of the two significantly enriched among the patients stratified by hospitalised status **(Table S8)**. Based on this data, rs12006793 may be an important marker for COVID-19 susceptibility and severity.

## DISCUSSION

Based on the hypothesis that a predisposing genetic background may contribute to the observed clinical variability of COVID-19, we set out to investigate whether variation in *ACE2* might modulate susceptibility to SARS-CoV-2 and influence severity. While large-scale GWAS have been conducted on COVID-19 patients,^33–36^ our study included the investigation of rare susceptibility alleles, which are often excluded in GWAS. Further, genetic variation in *ACE2* regulatory regions is a strong candidate for identifying differences in ACE2 activity. Yet, studies on genotype-dependent ACE2 expression in SARS-CoV-2 patients are currently lacking.

We systematically analysed *ACE2* polymorphisms associated with higher protein expression and coding-region variants between patients and controls, as well as individuals from different ethnicities. We provide evidence for differences in coronavirus S protein binding affinity between ACE2 variants in different ethnicities and eQTLs significantly associated with COVID-19 susceptibility.

Among 1,837 SARS-CoV-2 positive patients, six nonsynonymous *ACE2* variants (V316L, H345R, H401R, M455V, Y515N, and P737L) are not included in public databases, such as gnomAD, dbSNP and 1KGP3. Based on the functional analysis of a synthetic human ACE2 mutant library for RBD-binding affinity,^22^ p.Y515N reduces and p.K29R increases the receptor affinity for the viral S protein. Further, the top variants predicted to disrupt protein function based on the tools used in this study include p.P263S (rs200745906), p.H345R (novel), p.H378R (rs142984500), p.H401R (novel), p.Y515N (novel), and p.S692P (rs149039346). Some of these damaging ACE2 missense variants may be protective against SARS-CoV-2 infection, by either disrupting the proteolytic activation of SARS-CoV-2 or decreasing S-protein binding.

Two missense variants (K26R and N720D) were statistically more prevalent in the European population. We postulate that K26R increases the affinity of ACE2 for SARS-CoV-2, while N720D enhances TMPRSS2 activation and subsequent viral entry.

By performing structural modelling of mutations, we show that the H378R variant, which mutates one of the Zn binding histidines to arginine, could directly weaken the binding of catalytic metal atoms and reduce ACE2 peptidase activity. Further, H401R is located very close to the Zn binding pocket and could have a destabilising effect on the ACE2 structure.

Another potential explanation for different prognostic outcomes in these patients may involve ACE2 receptor expression levels. While the function of the rs199651576 T/C 5’UTR SNP remains unclear, it is possible that it is involved with the translational regulation of the ACE2 mRNA transcript. The splice site variant rs2285666 that was enriched in SARS-CoV-2 positive patients relative to 100KGP controls, and in hospitalised European patients relative to outpatients in intra-ethnic comparisons, increases ACE2 expression possibly via mRNA alternate splicing mechanisms.^24^ Studies have shown rs2285666 is correlated with hypertension, coronary heart disease and type 2 diabetes.^26,27^ This variant is more common among South Asians and East Asians and may be useful in predicting COVID-19 outcomes.

Twenty-three eQTLs for the *ACE2* gene were significantly different between patients and controls after multiple-comparison correction. Of these, ten with minor alleles that associated with higher tissue expression of ACE2 were associated with COVID-19 aetiology. Intriguingly, two eQTLs whose MAF associated with lower tissue expression were associated with reduced risk of testing positive for SARS-CoV-2. One of these polymorphisms (rs12006793) had a Cohen’s large effect of 0.91, suggesting it may be a useful marker for reduced susceptibility.

These eQTLs have considerable differences in allele frequencies among ethnicities within the 100KGP dataset **(Figure 4)**. Consistent with our findings, Cao et al.^6^ showed that the East Asian populations have much higher allele frequencies in the eQTL variants associated with higher ACE2 expression, suggesting population differences in response to SARS-CoV-2. In a study analysing single-cell RNA sequencing data of human lungs of eight donors, Zhao et al.^37^ showed that ACE2 was more abundantly expressed in type II alveolar epithelial cells of a male Asian donor compared to both Black and White donors.

Given that East Asians had a higher frequency of upregulating variants and lower frequency of downregulating variants compared to other populations, the resultant higher levels of ACE2 in this population may lead to higher COVID-19 susceptibility. Further, Africans showed a genetic predisposition for lower expression levels of ACE2, implicating the opposite. It is possible that the effect of individual eQTLs is too small to affect the receptor function of ACE2. However, genetic trait scores are increasingly used to make predictions of phenotype based on a cumulative contribution of genetic factors to a specific trait. Herein we highlight variants of significance in COVID-19 patients as a means of refining estimates of individual liability to disease **(Figure 5)**.

**Figure 5:**
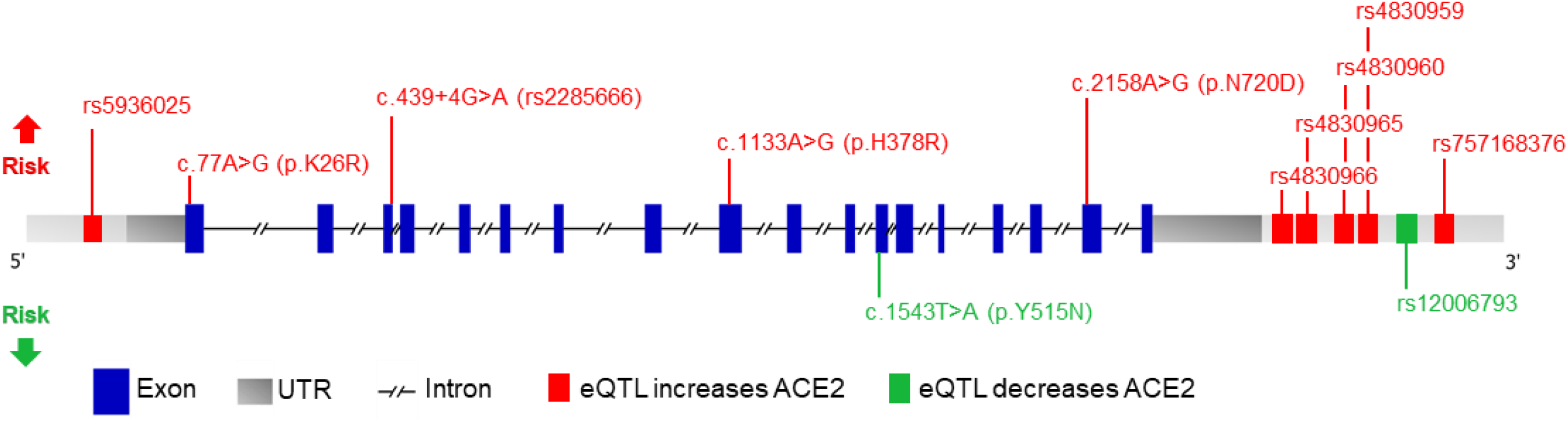
Schematic representation of the ACE2 gene with variants affecting risk. Genomic structure is based on the Ensembl canonical transcript (ENST00000252519.8), which has 18 exons, a transcript length of 3,339 bps, and a translation length of 805 residues. Exons are shown as blue boxes, the 5’ and 3’ UTRs are shown as grey boxes, and the horizontal black dashed line represents the introns. The eQTLs that increase and decrease ACE2 expression levels are depicted as green and red boxes, respectively. Only eQTLs with a Cohen’s d effect size > 0.2 are shown. Missense and splicing variants are shown with their CDS and protein position. The locations of the variants associated with increased and decreased COVID-19 susceptibility are shown above and below the gene, respectively.

The 100KGP dataset shows a high proportion of participants testing positive for SARS-CoV-2 and hospitalised compared to the rest of the population. This could partly be a reflection of increased interactions with the healthcare system, or that patients with pre-existing comorbidities are at increased risk of COVID-19. While this may not be a true representation of the UK population, studying at-risk individuals offers a unique opportunity to investigate variability in COVID-19 outcome. This may lead to improved prevention measures, triage strategies and clinical intervention.

In this study, we provide further insights into the potential role of genetics on SARS-CoV-2 infectivity and disease severity. We provide evidence of a genetic link between the *ACE2* genotype and COVID-19 disease severity and suggest that both eQTLs and coding variants may inform COVID-19 risk stratification.

## Data Availability

This research was made possible through access to the data and findings generated by the 100,000 Genomes Project.

## ACKNOWLEDGEMENTS

This research was made possible through access to the data and findings generated by the 100,000 Genomes Project. The 100,000 Genomes Project is managed by Genomics England Limited (a wholly owned company of the Department of Health and Social Care). The 100,000 Genomes Project is funded by the National Institute for Health Research and NHS England. The Wellcome Trust, Cancer Research UK and the Medical Research Council have also funded research infrastructure. The 100,000 Genomes Project uses data provided by patients and collected by the National Health Service as part of their care and support.

